# Validity of food portion size photographs among women in Nairobi, Kenya

**DOI:** 10.64898/2025.12.28.25343121

**Authors:** Ifrah Abdirashid Haji, Mikael Fogelholm, Hanna Mari Walsh, Noora Kanerva

**Author notes:** Correspondence: Noora Kanerva, Department of Food and Nutrition, POB 66, 00014 University of Helsinki, Finland.

## Abstract

**Background:** Validated food portion size photographs can increase accuracy of estimating portion sizes during dietary surveys. Our objective was to assess the validity of food portion size photographs in estimating portion sizes to be used in 24-hour dietary recall food consumption study.

**Methods:** We recruited two hundred and six women of reproductive age (13-45 years) residing in Roysambu sub-county in the northern parts of Nairobi City, Kenya. Eleven foods from the Photographic Food Atlas for Kenyan Adolescents (9-14 years) were examined. Participants were served pre-weighed portions. After eating, each participant was asked to estimate the amount of food they consumed, using the Food Atlas. Validity was assessed by calculating percentage of estimates within and outside of ±10% of consumed portion size, the mean percent difference between estimated and consumed portions, Spearman’s correlation coefficients, and Pearson’s chi-square test.

**Results:** The proportion of participants with estimates within ± 10% of the consumed portion size ranged between 15-65%. Mean differences between the consumed and estimated portion sizes varied -45% for stewed beans to +60% for watermelon. Generally, small portions were overestimated while large portions were underestimated. Correlation coefficients ranged from 0.12 to 0.77 and all the coefficients were statistically significant except for watermelon (p=0.22). Accuracy of estimations was not associated with participants’ age or educational level.

**Conclusions:** The validity of the tested food proportion size photographs was adequate for quantifications of most food items. However, the study’s findings also indicated that further improvement is needed before wider use in Kenya.

## Background

A growing number of low and middle-income countries, including Kenya, are facing the double burden of malnutrition [1–3], defined as the coexistence of undernutrition with overweight or obesity and dietary-related non-communicable diseases [4]. The causes of double burden of malnutrition have been attributed to various factors including marked transition in dietary patterns, a decrease in physical activity and insufficient access to healthy food options [3]. The changes in diet and physical inactivity may associate with an increase in non-communicable diseases (NCDs) in Kenya such as hypertension and diabetes [5]. Women are especially vulnerable to malnutrition due to their high nutritional requirements during pregnancy and lactation [6].

According to findings from the Kenya Demographic and Health Survey, girls aged 15-19 years were reported to be the most malnourished group among women of reproductive age with nearly 17% underweight while those between 40-49 years were the most overweight or obese (48%) [7]. Moreover, a recent cross-sectional study conducted in Nairobi City reported 18% prevalence of overweight (BMI z-score >1.0) and 7% prevalence of underweight (BMI z-score < -2.0) among boys and girls aged 9-14 years [8]. Furthermore, prevalence of overweight among adult women was (BMI >24.9kg/m^2^) 66 % and prevalence of underweight (< 18.5 kg/m^2^) 2%. Hence, valid assessment methods and accurate data of women’s dietary intake, in particular, are essential for nutrition status research and surveillance to inform public health policies and interventions.

In a resource-constrained context, such as Kenya, methods like the 24-hour dietary recall interview are preferred to assess dietary intake [9] as they impose little burden on respondents and do not require literacy compared to weighed food diaries [10]. A major source of error in dietary assessment in large-scale surveys is the assessment of portion size [11, 12]. Food photographs are useful tools for enhancing the accuracy of portion size estimations among different age groups, including adults [13, 14] as well as children and adolescents [15, 16].

Moreover, they are beneficial due to their adaptability to local conditions, low cost, portability [17] and, when compared to other methods, are widely regarded as the tool that best represents the actual food consumed [18]. Nonetheless, a recent systematic review highlighted that there was lack of validated estimation aids, emphasizing the need for more validation studies [12].

A photographic Food Atlas with 173 food items and dishes was developed in 2018 to evaluate portion sizes of Kenyan 9-14 year old boys and girls living in Nairobi [19]. Though the Food Atlas was developed initially to be used in a study involving for 9-14-year-olds, we were also going to use it when assessing women of reproductive age. Therefore, the present study aimed to assess the validity of selected food portion size photographs from the Food Atlas among a broader age group of women of reproductive age (13-45 years). As it has been for long known that individual’s sociodemographic characteristics affect reporting of dietary intake [20–23], the secondary aim was to assess the association of sociodemographic characteristics such as age and educational level with the accuracy of portion size estimation.

## Methods

### Participants and settings

The validation study was done in preparation for the InnoFoodAfrica (EU Horizon grant agreement No 862170) food consumption survey, which aimed to assess food intake and consumption habits among mothers and their infants (6-23 months) in Nairobi City and Chuka Town in Kenya using a 24-hour dietary recall interview. The validation study was undertaken in Kahawa West region of Nairobi County, within the sub-county of Roysambu in the northern parts of Nairobi County. The location was chosen strategically to capture and represent a range of low and middle socioeconomic status (SES) households, which corresponded to the SES groups for which the Food Atlas was designed. Additionally, the region was one of the two locations where the InnoFoodAfrica food consumption study would be conducted.

The study participants were women of reproductive age (13-45 years). The research team recruited them by word- of- mouth, through the distribution of information leaflets, and through the help of three local community health volunteers who were advertising the study in their dedicated areas. The inclusion criterion of the study were girls and women between the ages of 13 and 45, with no conditions preventing them from eating the foods served such as allergies or following a special diet, and that they were not ill at the time of the study. We aimed at having a sample covering the range important socio-demographic background variables of the InnoFoodAfrica food consumption study, mainly age and socio-economic status.

Based on earlier similar validation studies [16] we targeted to have at least 200 participants enrolled in the study. Thus, the recruitment process continued until the target of 200 girls and women consenting to participate in the study was reached. We estimated that having 200 participants would result in having an analytical sample of 30 participants for each of the estimated foods. The final number of participants varied between 40 (Sukuma wiki/collard greens and Tilapia fish) and 112 (sweet banana) participants. Based on post-hoc analysis for achieved power done by using G*Power software (version 3.1.9.7; Heinrich-Heine-Universität Düsseldorf, Düsseldorf, Germany), our lowest participant number (n = 40) gave us 0.98 power to detect correlation coefficient r = 0.57 with a = 0.05.

### Kenyan Photographic Food Atlas

The Photographic Food includes 173 common Kenyan foods and dishes [19] and it was developed by following a five-step theory [24]. Most of the food items in the Atlas were presented in three increasing portion sizes, A (small), B (medium), and C (large). For foods that come in clearly defined individual pieces or packages, for instance fruits, we did not always calculate the portion sizes according to this formula since they may have a different ratio between the portion sizes. However, having three portion sizes is perhaps the most common number of portions presented in this type of photograph series [11, 16, 25, 26]. Portion B was the average portion size in most of the foods, determined by calculating an average from all the weighed portions consumed by the adolescents during data collection. Meanwhile, portion A was half (0.5) of portion B while portion C was one and half times (1.5) bigger than portion B for most of the food items.

Assessing the validity of the entire photographic atlas (173 foods) against a real served and weighed portion was not possible. Therefore, we chose 11 foods for the present evaluation: *ugali* (stiff maize flour porridge), *chapati* (flat bread), rice, stewed beans, fried tilapia fish, stewed beef, *sukuma wiki* (collard greens), sweet banana, orange, pawpaw, and watermelon (Table 1). The selection of foods was based on the criteria that they all were very commonly consumed by Kenyan adults, and they were foods that contribute significantly to energy and/or nutrient intake in the Kenyan population [27–29].

**Table 1.** Description of food items included in the validation.

| Food category | Food items | Texture |
| --- | --- | --- |
| Cereals based foods | *Ugali* (stiff maize flour porridge) | Amorphous |
|  | *Chapati* (flat bread) | Solid |
|  | Rice | Amorphous |
| Legumes | Stewed beans | Amorphous |
| Fish and meat dishes | Fried tilapia fish | Solid |
|  | Stewed beef | Amorphous |
| Vegetables | *Sukuma wiki* (collard greens) | Amorphous |
| Fruits | Sweet banana | Whole fruit |
|  | Orange | Whole fruit |
|  | Pawpaw | Cut pieces |
|  | Watermelon | Cut pieces |

### Study design and implementation

The study team consisted of two research leaders and four trained research assistants (all with at least a bachelor’s degree in human nutrition). The data collection lasted 10 days, and a study chef prepared a different menu for lunch each day. The lunch included a combination of 4-5 of the 11 test foods presented in Supplement Table 1. Once participants arrived at the study location (a temporary lunch facility at the premises of a local church), they were given a brief oral presentation in addition to the written information and consent forms. Participants were informed they would be offered food and questioned about it after eating, but they were not told that they would be asked about the food portions they consumed. Data collection took part after signing the consent form. First, the participants were interviewed about their socio-demographic background, including questions about participants’ age, marital status, education level, occupation, and pregnancy status, adapted from the Kenya Demographic Health Survey questionnaire [7].

Portion sizes served to participants were weighed out into A, B and C portion sizes as depicted in the Food Atlas and randomly allocated to participants following a pre-established random list.

Food portions were weighed with a VON Hotpoint kitchen weighing scale with a 1g accuracy. Water was available for drinking with the meals. All food items constituting the meals were served on the same plate, and the (fruits) on a separate plate. The participants could choose their seat freely and there was no placing based on what portion sizes they got. However, they reported their consumption privately to an enumerator.

Approximately 5-8 minutes after the meal, each participant was directed into a separate area where they were interviewed. To avoid interviewer bias, the members of the study team who conducted the interview were not the same as those who served the food, and thus were unaware of the portion size served to the participants. Participants were asked to estimate the portion sizes with the help of the Food Atlas. Food photographs of the served foods from the Foot Atlas were shown to the participants following the dining session. The assessment began by asking the participant to indicate which of the photographs best represented the amount of food they were served, and how much of it she had consumed. The participants were told they could also choose between the photographs, choose fractions of a photograph portion or choose smaller than the smallest or larger than the largest depicted portion size. When participants reported that they did not consume all of the foods served to them, they were asked to describe their leftovers in spoonful or by using the photographs.

If any food remained, it was weighed and the weight was noted down, and the amount of food consumed was calculated as the difference between the amount initially served and the amount left. Leftover foods were separated before weighing them each separately. In contrast to the Food Atlas, fruits such as watermelon, sweet banana, and orange were weighed with their peels during the study; hence, the average portion of peel weight for each fruit was determined from 10 samples prior to the study to obtain the correct weight. As a result, the peel proportions were as calculated follows: watermelon 35%, sweet banana 30%, and orange 25% from the total weight of the fruit.

## Statistical analysis

Statistical analyses were carried out using IBM Statistical Package for the Social Sciences SPSS, version 26. Frequency distribution and percentages were used to describe the characteristics of the participants.

The validity of the estimated portion sizes against consumed (weighed) portions was evaluated using four different approach:

1. Percentage of participant estimates within and outside of ±10% of consumed portion size [16].
2. The differences between the estimated and consumed portion sizes of each food was calculated in two ways: as the difference in grams (estimated – consumed), followed by percentage difference [(estimated- consumed)/consumed] *100, means and 95% confidence intervals were calculated for both. A positive value indicated an ‘overestimation’ and a negative value an ‘underestimation’ of portion sizes.
3. Spearman’s rank correlation coefficient was calculated to assess the association between the estimated consumed and portion sizes.
4. Pearson’s chi-square test was used to determine the association between errors in food portion estimation by age and educational level.

The significance level for statistical analyses was set at 0.05 for all tests.

## Ethical approval

All procedures involving research study participants were approved by the Chuka University Ethics Review Committee as part of the Ethical clearance of the InnoFoodAfrica project, and permission to conduct the study in Nairobi County was obtained from the National Commission for Science, Technology, and Innovation (NACOSTI; license number NACOSTI/P/21/8890).

Participation in the study was voluntary. Researchers informed the participants about the objective of the study when they arrived at the study location (at the Kahawa West Church). After explaining the study information, the participants had possibility to ask further questions and clarifications. Written informed consent was obtained from all participants and their data was handled pseudonymized. For participants under the age of 18 years, we ensured they were accompanied by a parent or guardian to the study area who also gave their written consent.

Names of the participants were not recorded in the electric study forms, but each participant was given a study ID. Data was collected using laptops and kept in excel files secured with passwords. Signed consent forms were kept in locked room at Africa Harvest Biotech Foundation International’s office.

## Results

### Participant characteristics

The participant demographics are presented in Table 2. A total of 209 women took part in the study. One participant dropped out after signing the informed consent, and two had duplicate identification numbers, hence data were available for 206 participants. The largest age-group (25% of participants) was 25-29-year-olds. Single women made up 67% of the study participants, and 6% of the participants reported being pregnant at the time of the study. A total of 36% had completed secondary education, while only 6% had less than a primary education. Thirty per cent were unemployed, and 9% were students.

**Table 2.**
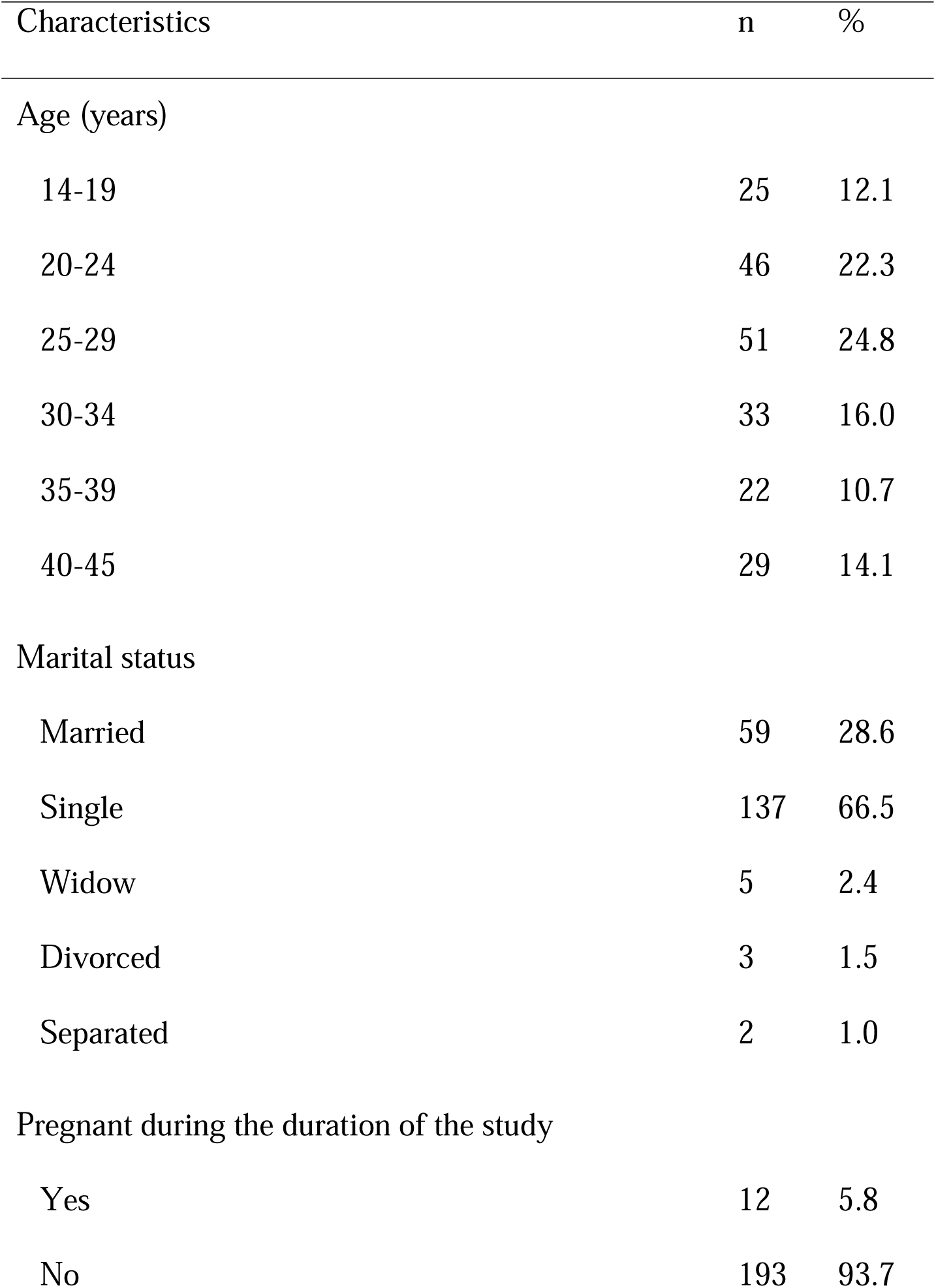
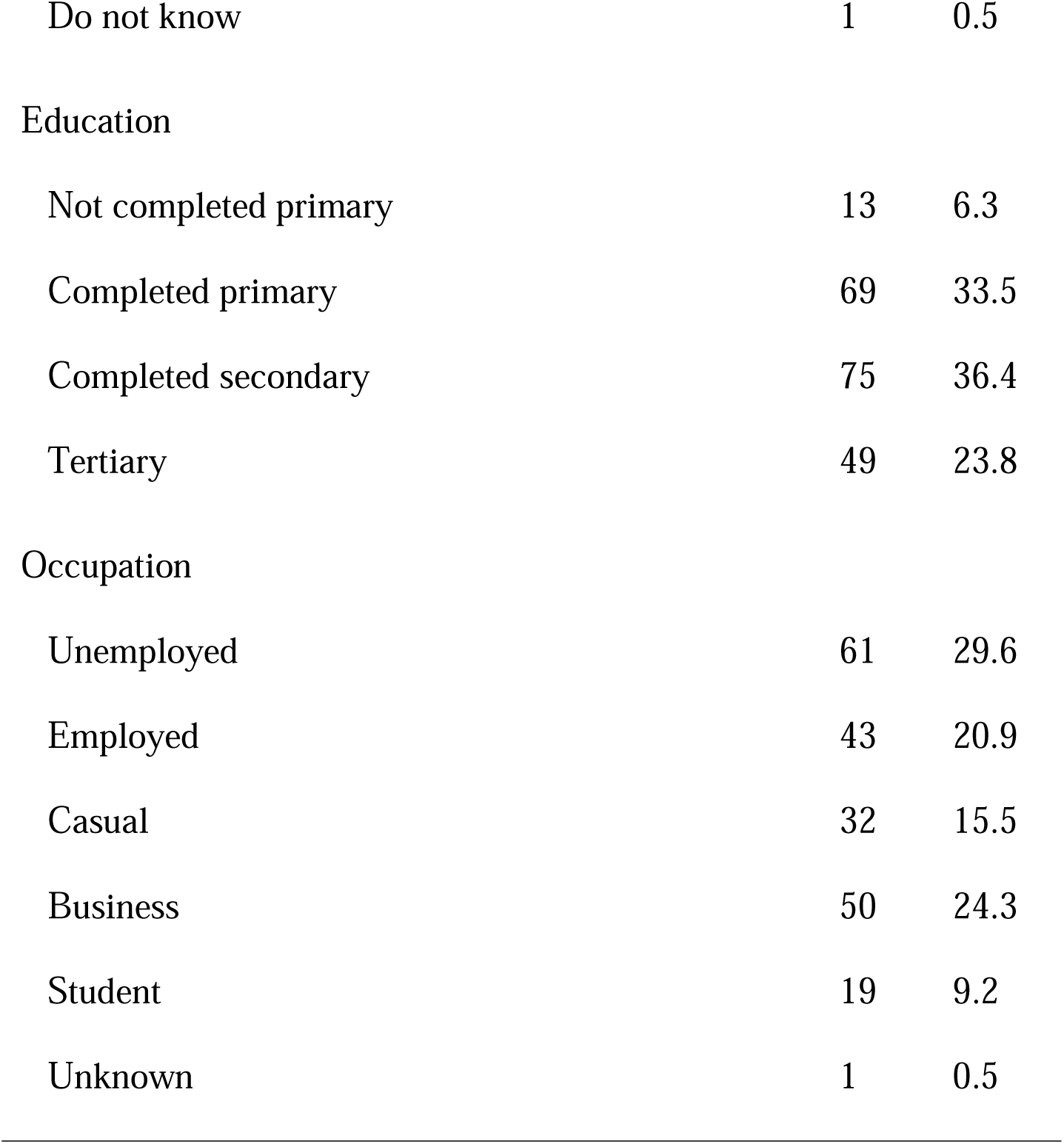
Participant characteristics: women of reproductive age (n = 206) participating in food portion size photograph validation, Kenya, 2021.

| Characteristics | n | % |
| --- | --- | --- |
| Age (years) |  |  |
| 14-19 | 25 | 12.1 |
| 20-24 | 46 | 22.3 |
| 25-29 | 51 | 24.8 |
| 30-34 | 33 | 16.0 |
| 35-39 | 22 | 10.7 |
| 40-45 | 29 | 14.1 |
| Marital status |  |  |
| Married | 59 | 28.6 |
| Single | 137 | 66.5 |
| Widow | 5 | 2.4 |
| Divorced | 3 | 1.5 |
| Separated | 2 | 1.0 |
| Pregnant during the duration of the study |  |  |
| Yes | 12 | 5.8 |
| No | 193 | 93.7 |
| Do not know | 1 | 0.5 |
| Education |  |  |
| Not completed primary | 13 | 6.3 |
| Completed primary | 69 | 33.5 |
| Completed secondary | 75 | 36.4 |
| Tertiary | 49 | 23.8 |
| Occupation |  |  |
| Unemployed | 61 | 29.6 |
| Employed | 43 | 20.9 |
| Casual | 32 | 15.5 |
| Business | 50 | 24.3 |
| Student | 19 | 9.2 |
| Unknown | 1 | 0.5 |

### Food portion size evaluations

The proportion of participants with estimates within ±10 % of the actual consumed portion size are presented in Table 3. Overall, for 4 out of the 11 food items more than half of the participants had estimated the portion size to be close the consumed portion size. These food items were, *chapati* (65%), *sukuma wiki (55%)*, rice (54%) and stewed beef (52%). In contrast, less than 20% of participants’ estimations were within ±10 % for sweet banana (18%) and watermelon (15%). For *ugali*, pawpaw, orange, tilapia and stewed beans, the proportion of estimates within ±10% ranged from 47% to 34%.

**Table 3.** Proportion of participants whose estimates were within ±10% of consumed portion size: women of reproductive age (n 206) participating in food portion size photograph validation, Kenya, 2021.

| Food item | Number of estimations | Estimates within $\pm 10\%$ of consumed portion size | | Underestimation more than -10% of consumed portion size | | Overestimation more than +10% of consumed portion size | |
| --- | --- | --- | --- | --- | --- | --- | --- |
|  | n | n | % | n | % | n | % |
| *Chapati* (flat-bread) | 57 | 37 | 64.9 | 16 | 28.1 | 4 | 7.0 |
| *Sukuma wiki* (collard greens) | 40 | 22 | 55.0 | 17 | 42.5 | 1 | 2.5 |
| Rice | 59 | 32 | 54.2 | 19 | 32.2 | 8 | 13.6 |
| Stewed beef | 115 | 60 | 52.2 | 50 | 43.5 | 5 | 4.3 |
| *Ugali* (stiff maize flour porridge) | 90 | 42 | 46.7 | 28 | 31.1 | 20 | 22.2 |
| Pawpaw | 95 | 43 | 45.3 | 20 | 21.1 | 32 | 33.7 |
| Orange | 88 | 39 | 44.3 | 27 | 30.7 | 22 | 25.0 |
| Tilapia | 40 | 17 | 42.5 | 20 | 50.0 | 3 | 7.5 |
| Stewed beans | 50 | 17 | 34.0 | 28 | 56.0 | 5 | 10.0 |
| Sweet banana | 112 | 20 | 17.8 | 62 | 55.4 | 30 | 26.8 |
| Watermelon | 105 | 16 | 15.2 | 31 | 29.5 | 58 | 55.2 |

The mean estimated consumed and portion sizes, as well as mean differences in grams, and mean percentage differences of the 11 foods tested are presented in Supplement Table 2. Mean differences for food items that were underestimated ranged from −1.4% for small portions of *chapati* to -45% for large portions of stewed beans. Mean differences for food items that were overestimated ranged from +1.1% for the small portions of stewed beef to +60% for the small portions of watermelon. Generally, small portions were overestimated while large portions were underestimated.

The Spearman’s rank correlation coefficients between the estimated and actual consumed portions ranged from 0.12 for watermelon to 0.77 for *chapati.* All of the coefficients were statistically significant, except for watermelon (p = 0.22; Table 4).

**Table 4.**
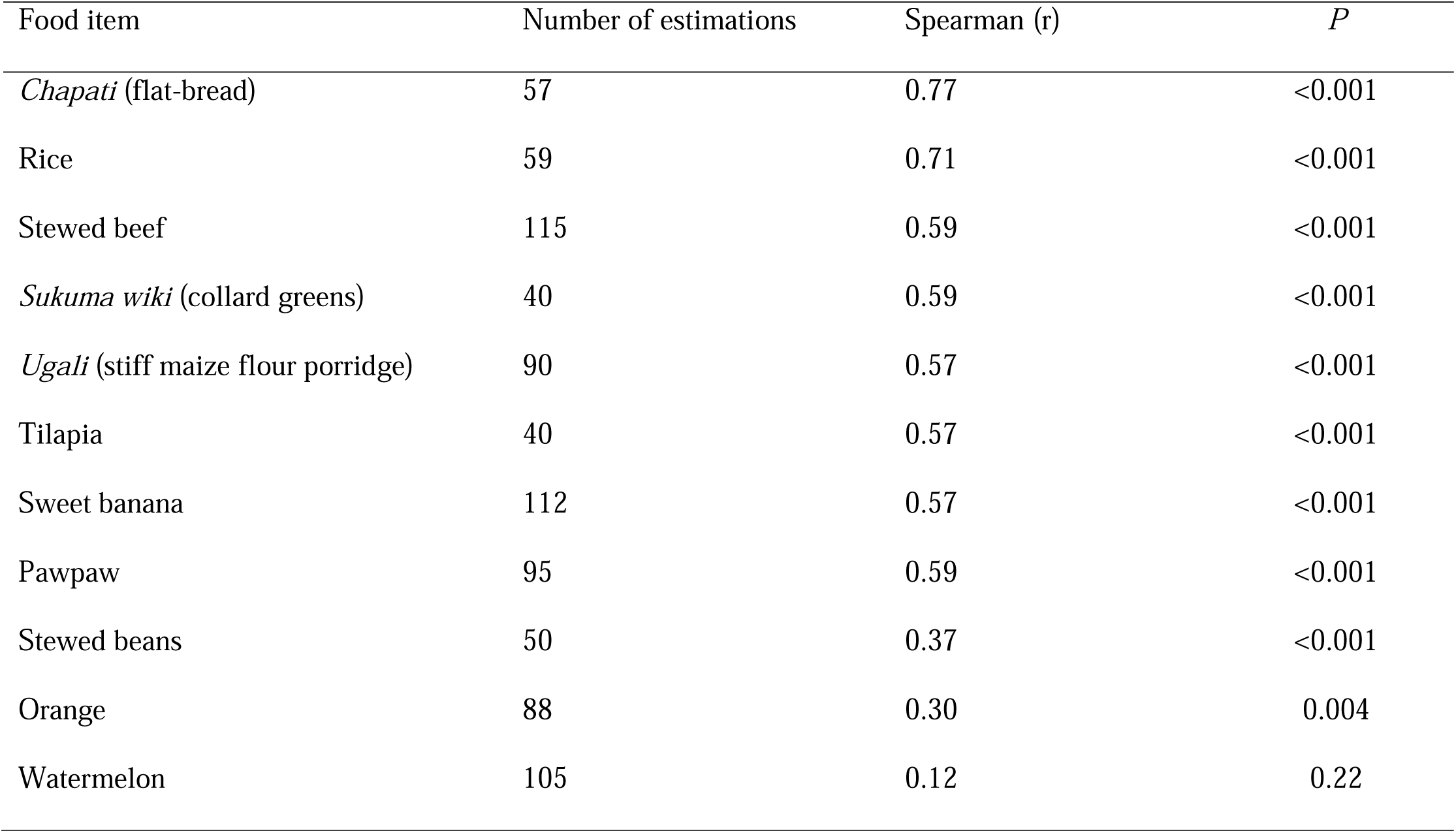
Spearman’s rank correlation coefficient between consumed and estimated portions: women of reproductive age (n = 206) participating in food portion size photograph validation, Kenya, 2021.

Results from the Pearson’s ^2^ analysis showed that the accuracy of portion size estimation did not differ across age groups or educational levels (Supplement Table 7). There were no statistically significant differences in the ability to estimate portion sizes using photographs between the age groups 14-24, 25-34, and 35-45, or between participants with higher (tertiary) education compared to those with other education.

## Discussion

Food portion size photographs can be useful for portion size measurement in population-based food consumption surveys in Kenya. In this study, we assessed the validity of food portion size photographs in a Food Atlas in estimating portion sizes for eleven commonly used foods among Nairobian women of reproductive age. Based on our results, food portion size photographs can be useful for portion size measurement in population- based food consumption survey in Kenya.

### Accuracy of portion size estimations

In the present study, the proportion of participants with close estimates, i.e., within ±10 % of the consumed portion size, ranged from 15% to 65% for different foods, with four food items having more than 50% accurate estimations. Other studies have mostly reported proportion of accurate estimates in the range of 50% or slightly higher [17, 25, 30, 31], and others lower [15, 16].

However, due to differences in study design, between-study comparability is challenging.

The mean differences between consumed and estimated portion sizes ranged from -45% for stewed beans to +60% for watermelon. When assessing the conceptualization skills of their participants, Korkalo et al. [16] found mean percentage differences between −19% for rice to +8% for shrimp sauce among adolescent girls, while Turconi et al. [11] reported mean percentage differences between -2.7 % for bread and 15.9 % for vegetables among their participants (6-60 years old). Participants in the latter study, however, self-served their own portions.

The accuracy between consumed and estimated portions ranged from a weak non-significant correlation for watermelon (*r* = 0.12) to a strong significant correlation for chapati (*r* = 0.77). Korkalo et al. [16] found correlations ranging from 0.46 for shrimp sauce to 0.73 for rice, whereas a Danish study reported correlations ranging from 0.18 for meat sauce to 0.89 for chocolate [15]. The latter study, however, differs in study design from the present study, making between-study comparability challenging.

The differences in portion size estimation accuracy in the present study can be attributed to many different factors such as food type (solid or amorphous), the portion sizes, the quality of photographs, the number of items per plate in the Food Atlas compared to the study meal, and leftover measurements. The results and potential sources of errors will be discussed further below, by first examining the characteristics of the Food Atlas, and then study design and methods.

### Characteristics of the Food Atlas

#### Food characteristics

It has been suggested that amorphous foods and meals, such as bean stew, which do not have a definite shape and that take the format of the container they are placed in, are associated with less precision [16, 17, 25, 31]. Contrary to what has been proposed, our participants were able to estimate the amorphous rice and stewed beef satisfactorily, whereas the assessment of stewed beans was clearly worse. Similarly, some solid foods were accurately assessed, while others were less accurately estimated. Other sources of error are likely to have played a larger role in our study.

#### Portion size photographs

The number of portion sizes in the Food Atlas should depict a wide enough range of portion sizes without being too arduous to go through and this can be a hard balance to strike. In our study, participants tended to overestimate the smallest portion sizes and underestimate the largest portion sizes. This is known as the ’flat slope phenomenon’ [17, 25]. This has been attributed to various factors, including whether participants were given the option of choosing a portion size smaller or larger than the one depicted in the photographs [17, 25]. Amougou et al. [25] asked participants in their study to estimate their portion sizes using one of the three depicted portions or the virtual portions placed in between the photographs. Despite being given the option, the authors reported that none of the participants chose a portion outside of the presented range.

Similarly, we gave our participants an option of selecting portions outside the presented range, but they never did. An increase in the number of portion options in the Food Atlas could have benefited the estimations so that participants would not automatically opt for the middle portion [30]. Another option would be to use additional letters between the photographs to depict virtual portion sizes [11, 25, 32]. This may have encouraged participants to report their portions more accurately [11].

Previous validation studies of food portion size photographs have shown that participants tend to have more difficulties in quantifying real food portions that differ from those in the photographs in terms of distribution on the plate, number of foods units and thickness of slices [14, 33, 34].

We noticed that photographs of *ugali* and stewed beans, were ambiguous in terms of the distribution area of food on the plates, making estimations problematic. In the case of stewed beans, for example, participants appeared to have difficulty in distinguishing between medium and large portions in the photographs, as the medium portion appeared to be larger due to the distribution of food on the plate. This resulted in the medium and large stewed bean portions being the most underestimated portion sizes, and overall stewed beans being the most underestimated food item. The same was also true for *ugali*, as participants had difficulty distinguishing between the small and medium portions. This may have contributed to the overestimation of small portions and underestimation of medium and large portions. Other authors have reported similar concerns with the intervals between portion sizes not being clear and noticeable resulting in a large proportion of the participants being classified in adjacent thirds of the distribution [16].

Our findings add to a growing body of literature that highlight the importance of presentation and shape of food items served in the food portion size photographs [33, 34]. It is crucial that each food portion is precise in term of the shape and outline, paying specific attention to sauce distribution. This is especially significant when we considering that our participants had to account for the portions of multiple food items on their plates, and any differences between the portions and what was served could have impacted estimation accuracy for all the food items.

### Methodological issues

#### Study design and participants

Food portion size photographs are assessed in relation to three cognitive skills of participants: perception, conceptualization and memory, which enable participants to relate a photographic depiction of a given amount of food to the amount of food actually consumed [35]. Perception refers to the ability of a participant to relate an amount of food present in reality to an amount depicted in a photograph, whereas conceptualization refers to the participant’s ability to develop a mental picture of a food portion not present and relate it to a photograph [35]. Memory refers to the participant’s ability to accurately recall the amount of food consumed [25]. The present study assessed participants’ conceptualization skills, which entailed serving them pre-weighed food items to consume and having them estimate the portion sizes using food portion size photographs without being able to see the portions in front of them (i.e., after eating the portion). The recall interviews were conducted a few minutes after meal consumption, hence the role of memory was minimized, though memory is necessary in the accuracy of conceptualization.

Nonetheless, in an actual 24-hour dietary recall, the interview is conducted the following day, and memory plays a more significant role. However, some authors have reported no significant differences in participants’ abilities to estimate food portion size 24 h after consuming the food compared to when the food was in view [13, 36].

In our study, age and educational level were not associated with accuracy of estimations, which is consistent with previous studies [11, 13, 16, 30]. However, although our total sample size was larger than typically recommended for validation studies [37], as well as larger than many previous similar studies [13, 14, 16], not all participants ate the same foods. This meant that the number of observations for some of the food items may have been insufficient across the different age and educational categories to detect any statistical significance.

The generalizability of our findings is limited by the fact that we used a convenience sample, which was not drawn at random from the target population in Nairobi. Nonetheless, we had a well-balanced sample in terms of age. Finally, our sample consisted of women of reproductive age from urban low and middle SES areas and households, and the results may be different in the very lowest and highest SES populations as well as populations residing in rural areas.

#### Weighing and the weight of food items

One limitation of the present study was the inability to match the weights of some of the food items to the weights in the Food Atlas. Our goal was to serve the amount of food as close as possible to its weight in the Food Atlas. However, due to the varying sizes of some of the foods that were available from the local market, such as tilapia, sweet bananas, pawpaw, and watermelons, this was not always possible.

In the case of fruits like watermelon and pawpaw, we had to serve a varying number of slices to obtain the exact weights, which was different from what was depicted in the photographs. This may have contributed to under-or overestimations of these two food items based on how they were presented. Other authors have reported similar lower accuracy when the food item served differed from the photographs [33, 34]. Due to diverse sizes of the fruits, it could be easier to simply provide photographs of various sized fruits rather than portion sizes, and during data collection participants can indicate the amount or fractions consumed using the photographs.

However, in real life situations people consume a variety of fruits that differ in sizes and shapes that do not always match those in the photographs, which allows us to get a more realistic picture of the estimation accuracy during dietary surveys.

Another source of weighing estimation error was related to leftovers, most common for the large portion sizes. For food items such as watermelon, tilapia, any edible flesh left was weighed and recorded as leftover. However, we noticed that when participants estimated their portions, despite being probed for leftovers, they may have regarded the flesh to be waste rather than leftover, and hence reported finishing their portions. This presents a challenge, particularly when assessing dietary intake of populations with diverse food cultures, such as Kenya, where what is regarded as leftover may have different meanings for different people, and any error in portion estimation would result in inaccurate picture of energy and nutrient intake.

In addition, a minor estimation error related to fruits, such as sweet banana, watermelon and oranges, could have resulted from weighing the peels. It was not always possible to weigh the proportion of peels for each fruit, as the leftover included both flesh and peels. As a result, the proportion of peel weight for each fruit was determined from ten samples before the study, and this proportion was assumed for each peel weight when calculating the amount of fruit consumed.

## Conclusions

We studied the validity of food portion size photographs from the photographic Food Atlas developed for Kenyan adolescents in assessing portion sizes of eleven commonly consumed food items in Kenya among women of reproductive age (13-45 years). The study assessed participants’ conceptualization skills by comparing consumed and estimated portions of the different food items.

The high proportion of participants who were able to estimate their portions using the food portion photographs for four of the food items including *chapati*, *sukuma wiki*, rice and stewed beef, indicates proper conceptualization of these staple foods. However, the accuracy was lower for the remaining seven food items. Correlations between consumed and estimated portions were significant for all foods items except for watermelon, and of similar magnitude as reported earlier.

Several characteristics of the Food Atlas may have contributed to assessment errors. Typical characteristics of the foods with the largest errors were the following: the number and size of food pieces on the plate was often different from what was shown in the Food Atlas; the photos may have been ambiguous and different portion sizes were not easy to differentiate; difficulties and potential inaccuracies in estimating and measuring the leftovers.

The current study is, to the best of our knowledge, the first to validate portion size photographs among women of reproductive age in Kenya. The present study bears international relevance as there is a lack of validated portion size estimation aids in Kenya and Sub-Saharan Africa in general. The tool appears to be reasonably accurate for estimating portion sizes of common foods in the Kenyan diet for Kenyan women of reproductive age. Further work to validate sets of food portion size photographs in different areas and population groups in Kenya and Africa is needed. This would allow for more accurate food intake assessment in Kenya and Africa leading to improved research and a better chance of addressing the double burden of malnutrition.

## Declarations

### Ethics approval and consent to participate

This study was conducted in accordance with the Declaration of Helsinki. It was approved by the Chuka University Ethics Review Committee as part of the Ethical clearance of the InnoFoodAfrica project, and permission to conduct the study in Nairobi County was obtained from the National Commission for Science, Technology, and Innovation (NACOSTI; license number NACOSTI/P/21/8890). All respondents provided written informed consent. Participants under the age of 18 years were accompanied by a parent or guardian to the study area who also gave their written consent.

### Consent for publication

Not applicable.

### Availability of data and materials

The data used for this manuscript are obtainable upon reasonable request.

### Authors’ contributions

IAH, NK, HMW, and MF designed the study. IAH, NK and MF participated in the data collection. IAH conducted the statistical analysis in close collaboration with NK and HMW. IAH and NK had the primary responsibility of drafting and finalizing the manuscript. HMW and MF reviewed the manuscript and approved the final version.

## Data Availability

All data produced in the present study are available upon reasonable request to the authors

https://kenfinedura.com/wp-content/uploads/2020/03/photographic-food-atlas-2-2-11-2018-1.pdf

## Acknowledgements

The authors thank the Kahawa West Full Gospel Church for providing facilities for this research. We are also thankful for Africa Harvest Biotech Foundation International for facilitating the study. We thank Ms. Wangari Kiragu for coordinating of the practical issues prior and during the study. Our gratitude goes to the field team members Ms. Sylvia Musinzi, Ms Sharlotte Awuor, and Mr. Felix Ondiek, and to the volunteers who took part in the study.

