## Supplementary tables 1-3 for "Validity of food portion size photographs among women in Nairobi, Kenya"

Supplement Table 1. Menu of the food items served to study participants.

| Week 1 | Week 2 |
| --- | --- |
| Monday:  *Ugali*, beef stew + orange and watermelon as dessert | Monday:  *Ugali* and beef stew + orange and melon as dessert |
| Tuesday:  *Chapat*i and bean stew + sweet banana and pawpaw as dessert | Tuesday:  Chapati and bean stew + sweet banana and pawpaw as dessert |
| Wednesday:  White rice and bean stew + sweet banana and pawpaw as dessert | Wednesday:  *Chapati* and beef stew + sweet banana and pawpaw as dessert |
| Thursday:  *Ugali*, fried tilapia and *Sukuma wiki* + orange and watermelon as dessert | Thursday:  *Ugali*, fried tilapia and *Sukuma wiki* + orange and watermelon as dessert |
| Friday:  White rice and beef stew + sweet banana and pawpaw as dessert |  |
| Saturday:  White rice and beef stew+ watermelon and sweet banana as dessert |  |

Supplement Table 2. Mean consumed and estimated portion sizes, mean difference in grams, mean percentage difference: women of reproductive age (n = 206) participating in food portion size photograph validation, Kenya, 2021.

| Food item | Portion size |  | Consumed portion size (g) | | Estimated portion size (g) | | Difference (g) | | % Difference | |
| --- | --- | --- | --- | --- | --- | --- | --- | --- | --- | --- |
|  |  | n | Mean | SD | Mean | SD | Mean | 95 % CI | Mean | 95 % CI |
| *Ugali* (stiff maize flour porridge) | A  B  C | 34  35  21 | 173.1  336.8  366.4 | 37.8  63.1  148.6 | 204.3  316.8  273.6 | 66.1  129.4  91.2 | 31.2  -19.9  -92.7 | 6.8, 55.5  -58.5, 18.7  -143.7, -41.7 | 29.5  -4.3  -4.6 | 4.0, 178.0  -18.5, 79.7  -36.0, 139.4 |
| Rice | A  B  C | 21  21  17 | 93.9  199.2  279.2 | 17.3  7.8  51.0 | 98.57  164.57  212.47 | 8.6  59.8  81.5 | 4.6  -34.6  -66.7 | 0.2, 9.0  -60.2, -9.0  -103.5, -29.9 | 7.72  -17.3  -23.1 | 0.6, 39.9  -30.1, 41.2  -36.0, 30.4 |
| *Chapati* (flat-bread) | A  B  C | 18  22  57 | 54.7  107.7  145.5 | 5.33  6.86  30.2 | 53.5  105.7  127.4 | 0  27.9  30.7 | -1.2  -1.9  -18.1 | -3.6, 1.2  -13.9, 10.0  -32.3, -3.84 | -1.4  -1.5  -10.8 | -9.8, 33.9  -12.5, 50.4  -19.89, 26.5 |
| Beef stew | A  B  C | 40  47  28 | 56.8  110.4  173.5 | 3.3  5.4  79.9 | 57.5  82.9  107.5 | 8.8  30.1  38.6 | 0.4  -27.5  -66.0 | -2.3, 3.2  -36.1, -18.9  -93.8, -38.1 | 1.12  -24.9  -34.3 | -3.8, 32.5  -32.6, 28.0  -43.3, 13.2 |
| Bean stew | A  B  C | 19  16  15 | 71.0  140.8  189.1 | 5.7  6.1  49.5 | 73.0  80.1  102.9 | 0  22.8  53.0 | 1.9  -60.7  -86.2 | -0.6, 4.52  -72.4, -49.0  -111.7, -60.6 | 3.4  -42.9  -45.4 | -0.6, 21.1  -51.2, -10.1  -57.5, 1.0 |
| *Sukuma wiki* (collard greens) | A  B  C | 14  15  12 | 72.6  141.7  181.6 | 1.7  2.1  44.1 | 75.3  107.3  119.4 | 19.4  36.1  38.3 | 2.6  -34.4  -62.2 | -7.9, 13.3  -52.5, -16.2  -81.8, -42.6 | 4.04  -24.3  -32.7 | -11.1, 60.8  -37.1, 25.4  -44.6, 8.4 |
| Tilapia | A  B  C | 13  15  12 | 120.7  160.8  258.0 | 23.87  12.42  51.75 | 86.85  137.2  202.5 | 27.8  23.4  59.0 | -33.9  -23.6  -55.5 | -16.4, -51.4  -8.0, -39.3  -4.0, -107.0 | -26.6  13.8  -17.2 | -40.8, 24.4  4.9, 48.2  -34.7, 43.2 |
| Orange | A  B  C | 34  35  21 | 114.2  123.3  148.0 | 13.0  37.4  42.1 | 115.6  127.5  118.0 | 31.4  29.2  33.6 | 1.43  1.17  -26.0 | -9.3, 12.2  -11.0, 13.3  -39.0, -12.98 | 2.0  44.2  -15.5 | 59.9, -7.8  546.6, -40.6  27.3, -24.8 |
| Sweet banana | A  B  C | 39  43  34 | 52.1  94.8  124.9 | 19.4  29.2  50.4 | 57.0  72.3  97.5 | 25.1  28.2  42.5 | 4.9  -22.5  -27.5 | -4.6, 14.3  -33.0, -12.1  -47.3, -7.7 | 27.8  -19.8  -2.5 | -11.0, 66.6  -30.1, -9.5  -36.8, 31.8 |
| Pawpaw | A  B  C | 33  37  29 | 75.8  141.0  198.5 | 26.9  45.7  99.3 | 99.6  154.6  186.8 | 54.8  63.6  105.1 | 23.9  13.6  -11.7 | 5.9, 41.8  -7.9, 35.1  -32.2, 8.8 | 28.4  26.4  1.9 | 6.8, 50.1  -2.1, 54.8  -16.7, 20.5 |
| Watermelon | A  B  C | 39  41  26 | 98.7  150.2  193.9 | 24.0  34.6  36.0 | 150.4  172.3  176.1 | 45.9  53.0  61.8 | 51.7  22.5  -17.8 | 36.3, 67.1  1.9, 43.1  -42.6, 7.0 | 59.6  36.6  -6.7 | 36.6, 73.9  -2.9, 76.1  -20.4, 7.1 |

SD= standard deviation.

Difference (g) = estimated –consumed.

% Difference= (estimated-consumed)/consumed X 100; (+) = overestimation; (-) = underestimation.

Supplement Table 3. Portion size accuracy by different age and educational level categories: women of reproductive age (n = 206) participating in food portion size photograph validation, Kenya, 2021.

|  |  | Within 10% | | Outside 10% | | P-value* |
| --- | --- | --- | --- | --- | --- | --- |
| Food item | Demographic characteristics | *n* | *%* | *n* | *%* | 0.172 |
| *Ugali* (stiff maize flour porridge) | Age, y  14-24  25-34  35-45 | 12  14  16 | 44.4  37.8  61.5 | 15  23  10 | 55.6  62.2  38.5 |  |
|  | Education  Primary  Secondary  Tertiary | 17  12  13 | 48.6  40.0  52.0 | 18  18  12 | 51.4  60.0  48.0 | 0.645 |
| Rice | Age, y  14-24  25-34  35-45 | 11  13  8 | 39.3  68.4  66.7 | 17  6  4 | 60.7  31.6  33.3 | 0.090 |
|  | Education  Primary  Secondary  Tertiary | 13  14  5 | 56.5  66.7  33.3 | 10  7  10 | 43.5  33.3  66.7 | 0.136 |
| *Chapati* (flat- bread) | Age, y  14-24  25-34  35-45 | 10  19  8 | 62.5  67.9  61.5 | 6  9  5 | 37.5  32.1  38.5 | 0.891 |
|  | Education  Primary  Secondary  Tertiary | 14  17  6 | 58.3  70.5  66.7 | 10  7  3 | 41.7  29.2  33.3 | 0.658 |
| Stewed beef | Age, y  14-24  25-34  35-45 | 13  30  17 | 39.4  60.0  53.1 | 20  20  15 | 60.6  40.0  46.9 | 0.183 |
|  | Education  Primary  Secondary  Tertiary | 26  19  15 | 45.6  52.8  68.2 | 31  17  7 | 54.4  47.2  31.8 | 0.197 |
| Stewed beans | Age, y  14-24  25-34  35-45 | 7  8  2 | 31.8  40.0  25.0 | 15  12  6 | 68.2  60.0  75.0 | 0.721 |
|  | Education  Primary  Secondary  Tertiary | 2  9  6 | 15.4  45.0  35.3 | 11  11  11 | 84.6  55.0  64.7 | 0.212 |
| *Sukuma wiki* (collard green) | Age, y  14-24  25-34  35-45 | 6  9  7 | 40.0  64.3  63.6 | 9  5  4 | 60.0  35.7  36.4 | 0.336 |
|  | Education  Primary  Secondary  Tertiary | 8  8  6 | 72.7  42.1  60.0 | 3  11  4 | 27.3  57.9  40.0 | 0.251 |
| Tilapia | Age, y  14-24  25-34  35-45 | 5  9  3 | 33.3  64.3  27.3 | 10  5  8 | 66.7  35.7  72.7 | 0.118 |
|  | Education  Primary  Secondary  Tertiary | 5  7  5 | 45.5  36.8  50.0 | 6  12  5 | 54.5  63.2  50.0 | 0.772 |
| Orange | Age, y  14-24  25-34  35-45 | 16  9  14 | 47.1  30.0  58.3 | 18  21  10 | 52.9  70.0  41.7 | 0.657 |
|  | Education  Primary  Secondary  Tertiary | 16  9  14 | 47.1  30.0  58.3 | 18  21  10 | 52.9  70.0  41.7 | 0.151 |
| Sweet banana | Age, y  14-24  25-34  35-45 | 10  6  4 | 23.3  13.3  16.7 | 33  30  20 | 76.7  86.7  83.3 | 0.471 |
|  | Education  Primary  Secondary  Tertiary | 9  6  5 | 20.0  14.0  20.8 | 36  37  19 | 80.0  86.0  79.2 | 0.693 |
| Pawpaw | Age, y  14-24  25-34  35-45 | 13  18  12 | 40.6  40.9  63.2 | 19  26  7 | 59.4  59.1  36.8 | 0.215 |
|  | Education  Primary  Secondary  Tertiary | 19  18  6 | 59.4  43.9  27.3 | 13  23  16 | 40.6  56.1  72.7 | 0.061 |
| Watermelon | Age, y  14-24  25-34  35-45 | 5  5  6 | 14.3  13.2  18.8 | 30  33  26 | 85.7  86.8  81.3 | 0.796 |
|  | Education  Primary  Secondary  Tertiary | 8  5  3 | 16.7  16.1  11.5 | 40  26  23 | 83.3  83.9  88.5 | 0.831 |

* P-value is from X^2^- test.
